# Deep profiling of multiple ischemic lesions in a large, multi-center cohort: Frequency, spatial distribution, and associations to clinical characteristics

**DOI:** 10.1101/2022.07.01.22277062

**Authors:** Anna K. Bonkhoff, Teresa Ullberg, Martin Bretzner, Sungmin Hong, Markus D. Schirmer, Robert W. Regenhardt, Kathleen L. Donahue, Marco J. Nardin, Adrian V. Dalca, Anne-Katrin Giese, Mark R. Etherton, Brandon L. Hancock, Steven J. T. Mocking, Elissa C. McIntosh, John Attia, John W. Cole, Amanda Donatti, Christoph J. Griessenauer, Laura Heitsch, Lukas Holmegaard, Katarina Jood, Jordi Jimenez-Conde, Steven J. Kittner, Robin Lemmens, Christopher R. Levi, Caitrin W. McDonough, James F. Meschia, Chia-Ling Phuah, Stefan Ropele, Jonathan Rosand, Jaume Roquer, Tatjana Rundek, Ralph L. Sacco, Reinhold Schmidt, Pankaj Sharma, Agnieszka Slowik, Alessandro Sousa, Tara M. Stanne, Daniel Strbian, Turgut Tatlisumak, Vincent Thijs, Achala Vagal, Daniel Woo, Ramin Zand, Patrick F. McArdle, Bradford B. Worrall, Christina Jern, Arne G. Lindgren, Jane Maguire, Ona Wu, Petrea Frid, Natalia S. Rost, Johan Wasselius, the MRI-GENIE and GISCOME Investigators and the International Stroke Genetics Consortium

## Abstract

**Background:** A substantial number of patients with acute ischemic stroke (AIS) experience multiple acute lesions (MAL). However, the spatial distribution and clinical implications of such MAL are incompletely understood.

**Methods:** Analyses relied upon imaging and clinical data of patients with AIS from the international MRI-GENIE study. Initially, we systematically evaluated the occurrences of MAL within (i) one and, (ii) several vascular supply territories. Associations between the presence of MAL and important characteristics, such as DWI lesion volume, NIHSS-based acute stroke severity, and long-term functional outcome were subsequently determined. The interaction effect between single and multiple lesion status and DWI lesion volume was estimated by means of Bayesian regression modeling for stroke severity and functional outcome.

**Results:** We analyzed 2,466 patients [age: 63.4±14.8, 39% women], 49.7% of which presented with single lesions. Another 37.4% experienced MAL in a single vascular territory, while 12.9% featured lesions in several territories. Within most territories (anterior, middle, and posterior cerebral artery, cerebellar), multiple lesions occurred as frequently as single lesions (ratio ∼1:1). Only the brainstem region comprised fewer patients with multiple lesions (ratio ∼1:4). Patients with MAL presented with a significantly higher DWI lesion volume and acute NIHSS (7.7ml vs. 1.7ml and 4 vs. 3, *p*_*FDR*_≤0.001). In contrast, patients with a single lesion were characterized by a significantly higher WMH burden (6.1ml versus 5.3ml, *p*_*FDR*_=0.048). Functional outcome did not differ significantly between patients with single versus multiple lesions. Bayesian analyses suggested that the association between DWI lesion volume and stroke severity between single and multiple lesions was the same in case of anterior circulation stroke. In the case of posterior circulation stroke, DWI lesion volume was linked to a higher acute NIHSS only among those with multiple lesions.

**Conclusions:** Multiple lesions, especially those within one vascular territory, occurred more frequently than previously reported. Overall, multiple lesions were distinctly linked to a higher acute stroke severity, a higher DWI lesion volume and lower WMH lesion volume. In posterior circulation stroke, lesion volume was linked to a higher stroke severity in multiple lesions only.

## Introduction

The stroke field has seen substantial advancements in recent years, particularly due to robust clinical trials that have impacted treatment pathways.^1^ Studies designed to improve our understanding of which lesion metrics are important for stroke outcomes can contribute to such progress in complementary ways, and support novel therapeutical approaches.

Neuroimaging, as a core element of modern acute stroke management,^2,3^ has been unparalleled in its capacity to derive detailed lesion characteristics. Especially after the introduction of elaborate imaging modalities, such as diffusion-weighted imaging (DWI), it became possible to reliably disentangle the acute versus chronic, lacunar versus non-lacunar, and single versus multiple nature of ischemic lesions, to name just a few examples. While all these lesion characteristics deserve further exploration, given their respective links to stroke mechanisms and outcomes, this present work investigates single versus multiple lesions.

An early computed tomography (CT)-based study estimated the frequency of multiple lesions to be ∼2%.^4^ Later magnetic resonance imaging (MRI)-focused re-evaluations, however suggested a frequency in the range of 10-30%.^5,6,7,8,9, 10,11,12,13^ These more recent estimates indicate that multiple lesions are a common and clinically relevant phenomenon. Concurrently, accumulating evidence suggests associations between the presence of multiple lesions and specific stroke etiologies, such as large-artery atherosclerosis^7,8^ and cardioembolism.^12^ In addition, multiple stroke lesions have also been linked to a higher initial stroke severity,^14^ higher risk of death^8^ and stroke recurrence.^15^

Lesion volume has long been known to explain substantial variability in stroke outcomes,^16,17,18^ positioning it as a clinically important prognostic marker. Conceivably, a higher lesion volume in the case of multiple lesions could readily explain the association with less favorable outcomes. However, the links between single and multiple lesion status, lesion volume and stroke outcomes have not been assessed thoroughly. This omission may, to a large extent, be due to the frequent unavailability of individual lesion volume information in large stroke datasets. Altogether, the in-depth investigation of lesion volume interaction effects with other lesion characteristics may be particularly important, as more and more studies indicate that the association between lesion volume and outcomes is more complex than initially believed. The links between lesion volume and long-term functional outcomes were, for example, shown to differ for small and large lesions.^19^ Further work indicated that only a fraction of the endovascular treatment benefit could be traced back to the reduction in lesion volume.^20^

We here build upon a large, uniquely well-phenotyped imaging dataset of patients with acute ischemic stroke (AIS) originating from the international, multi-site MRI–Genetics Interface Exploration (MRI-GENIE) study^21^ to i) radiologically phenotype multiple versus single ischemic lesions and ii) assess the effects between multiple lesions and lesion volume. We aimed to evaluate the hypothesis that lesion volume relates to stroke outcomes in varying ways depending on multiple versus single lesion status. Furthermore, the unique availability of i) a large dataset of multimodal clinical MRI scans and clinical information, ii) extensive automated segmentations of acute infarct lesions and white matter hyperintensity (WMH) lesions in combination with iii) comprehensive manual scan evaluations by neuroradiologist experts^22^ allowed us to perform analyses with an unprecedented level of detail.

## Methods

### Stroke patient sample

The present study relied on data of patients with AIS gathered for the international MRI– Genetics Interface Exploration (MRI-GENIE) study,^21^ which, itself, was based upon the Stroke Genetics Network (SiGN) collaboration.^23^ In brief, this study’s primary aim was to facilitate the genetic analysis of acute and chronic cerebrovascular neuroimaging phenotypes with an emphasis on creating a large database of acute and well-characterized MRI scans. While MRI-GENIE recruited 3,301 patients overall, we here focused on patients with complete radiological reports and discernible acute infarct diffusion-weighted imaging (DWI) lesions (n=2,468). Patients gave written informed consent in accordance with the Declaration of Helsinki. The study protocol was approved by Massachusetts General Hospital’s Institutional Review Board (Protocol #: 2001P001186 and 2003P000836).

### Neuroimaging data and structured reporting tool

MRI-GENIE patients underwent acute MRI examinations, with most examinations occurring within the first 48 hours of hospital admission and featuring DWI, as well as FLAIR sequences. In view of the multi-site character of the study, a variety of imaging parameters have been employed, a comprehensive overview is given in the **supplementary materials**. Two board-certified neuroradiologists (J.W. and M.D.) manually reviewed all individual scans and captured detailed information on lesion location (e.g., side, vascular territory) and further lesion characteristics (e.g., cortical/subcortical, single/multiple, lacunar/non-lacunar lesions for supratentorial strokes). An ischemic injury was classified as “multiple ischemic lesions stroke” if there were either multiple lesions within any of the predefined anatomical areas, or if there were distinct lesions in more than one of the predefined anatomical areas. Vascular territories encompassed the anterior cerebral artery (ACA), middle cerebral artery (MCA), posterior cerebral artery (PCA) and vertebrobasilar territory (cerebellum and brainstem), each coded separately for the left and right hemispheres. Importantly, lesions did not have to occur in multiple vascular territories to be considered as multiple lesions; i.e., we also assigned the multiple status within one vascular territory. It must be noted, however, that the borders are not distinct between vascular territories and substantial variation exists between individuals, therefore the association of lesions to vascular territories is somewhat subjective for such border-zone areas. An exhaustive description of the structured reporting tool used can be found in Drake and colleagues.^22^ Furthermore, DWI lesion volume, as well as WMH burden were estimated from automatically generated segmentations of DWI-defined stroke lesions^24^ and FLAIR-defined WM lesions, respectively.^25^

### Clinical data

Sociodemographic and clinical data comprised information on age, sex, stroke severity, stroke etiology, and comorbidities/cardiovascular risk factors. Stroke etiology was captured via the causative classification of stroke system (CCS),^26^ stroke severity was measured via the National Institutes of Health Stroke Scale during the acute hospital stay (NIHSS, 0-42, 0: no measured deficits, 42: maximum stroke severity). Functional outcomes were captured via the modified Rankin Scale (mRS, 0: no symptoms at all, 6: death) at 3-6 months. Comorbidities included hypertension, coronary artery disease, diabetes mellitus, atrial fibrillation, history of smoking and prior stroke.

### Descriptive statistics and group comparisons: Occurrence, spatial distribution, and clinical effects of single and multiple lesions

Our primary focus was the characterization of multiple ischemic lesions, i.e., their frequency of occurrence, spatial predilection, and associations to clinical factors and stroke outcomes. Multiple lesions were defined as >1 ischemic lesion, which could be located either within one vascular territory or multiple vascular territories. Patients were categorized according to the number of lesions and the number of involved vascular territories, i.e., single lesion in a single vascular territory, multiple lesions in a single vascular territory, two single lesions in two vascular territories, multiple lesions in two vascular territories, and so forth. Subsequently, we compared the frequency of single versus multiple lesions within each vascular territory. These descriptive statistics are reported as means and associated standard deviations (SDs). Associations of single versus multiple lesion status with clinical characteristics were evaluated via two-sample t-tests or Fisher’s exact tests as appropriate. The level of significance was set to *p*<0.05 after correction for multiple comparisons.

### Bayesian hierarchical regression: Interaction effects of multiple lesions and lesion volume with respect to functional outcome

In more granular analyses, we scrutinized the links between lesion volume and single versus multiple lesion status (as the exposures of interest) and stroke severity and unfavorable functional outcome (mRS>2) (as the outcomes of interest). These analyses were motivated by the hypothesis that patients with multiple stroke lesions would experience more severe strokes despite similar lesion volumes. Initially, we performed analyses for all stroke patients with available stroke severity and functional outcome data, and subsequently stratified for anterior versus posterior strokes. We excluded patients with lacunar stroke in the analysis of anterior circulation stroke patients, given that they per definition exclude multiple lesions. Lacunar stroke was defined as a single subcortical supratentorial lesion smaller than 1.5cm. The same analysis without the exclusion of patients with lacunar stroke is included in our **supplementary materials**. We explained acute stroke severity by means of Bayesian hierarchical *linear* regression. Similarly, we employed Bayesian hierarchical logistic regression to explain 3-to-6-months unfavorable functional outcomes. Log-transformed total DWI lesion volume represented the input to explain stroke severity, i.e., the acute NIHSS score, or unfavorable functional outcomes, i.e., mRS>2, as the output. Single versus multiple lesion status was integrated via the hierarchical structure of our model. In this way, we obtained an estimate of the association of DWI lesion volume with stroke severity separately for those patients with a single versus those with multiple lesions (c.f., **supplementary materials** for model specifications). As in previous work,^27,28^ we determined whether group estimates substantially differed via checking the overlap with zero for the *difference distributions* of posteriors (single – multiple lesions).

### Data availability and coding environment

Data can be made available to researchers for the purpose of reproducing the here reported results, pending the permission for data sharing by Massachusetts General Hospital’s institutional review board. Analyses were implemented in Python 3.7, hierarchical models relied on pymc3.^29^

## Results

We included a total of 2,466 MRI-GENIE patients with available MRI examinations and visible acute DWI stroke lesions in this study (mean age (standard deviation (SD)): 63.4 (14.8), 39.0% women, **Table 1**).

**Table 1.**
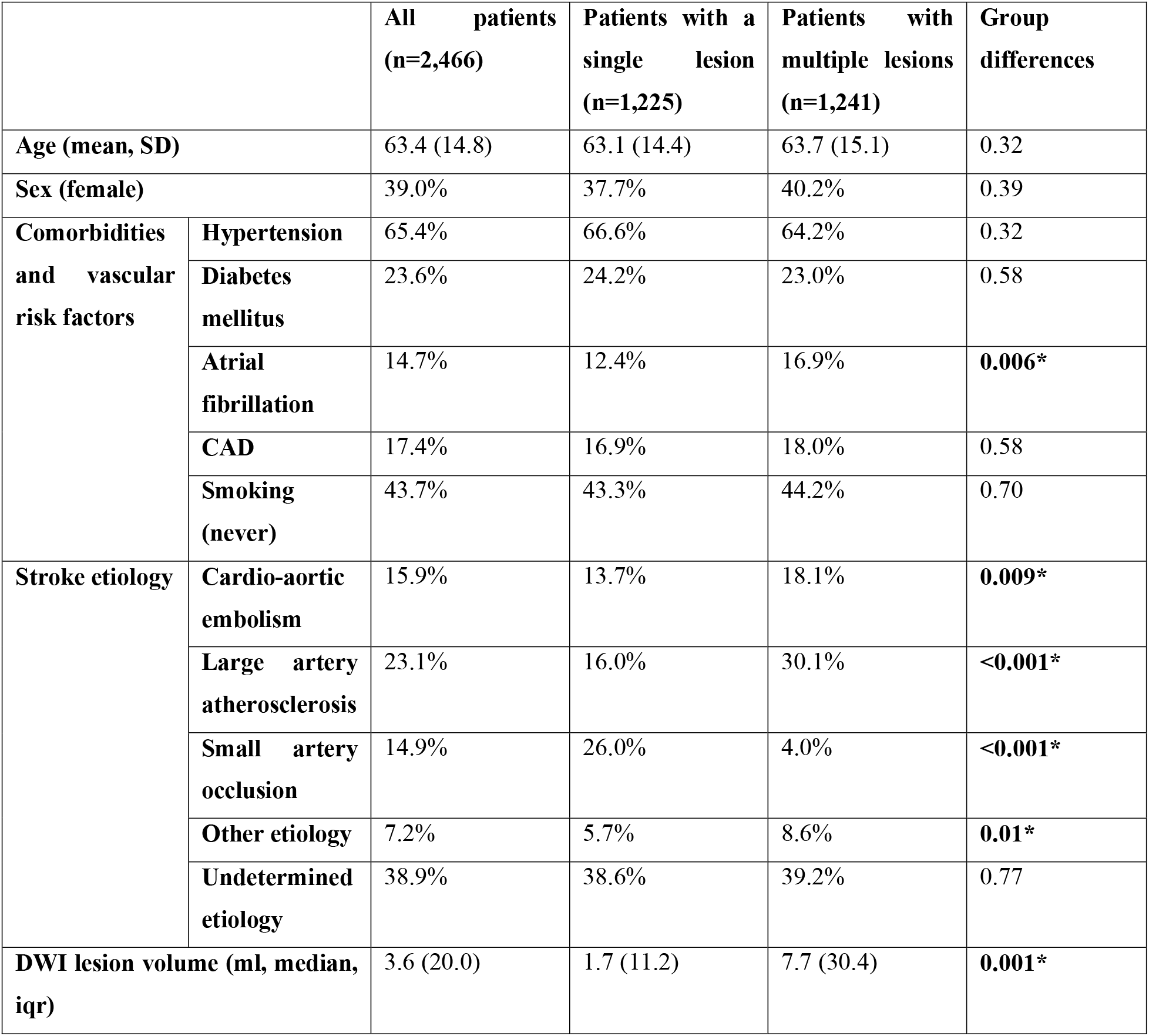
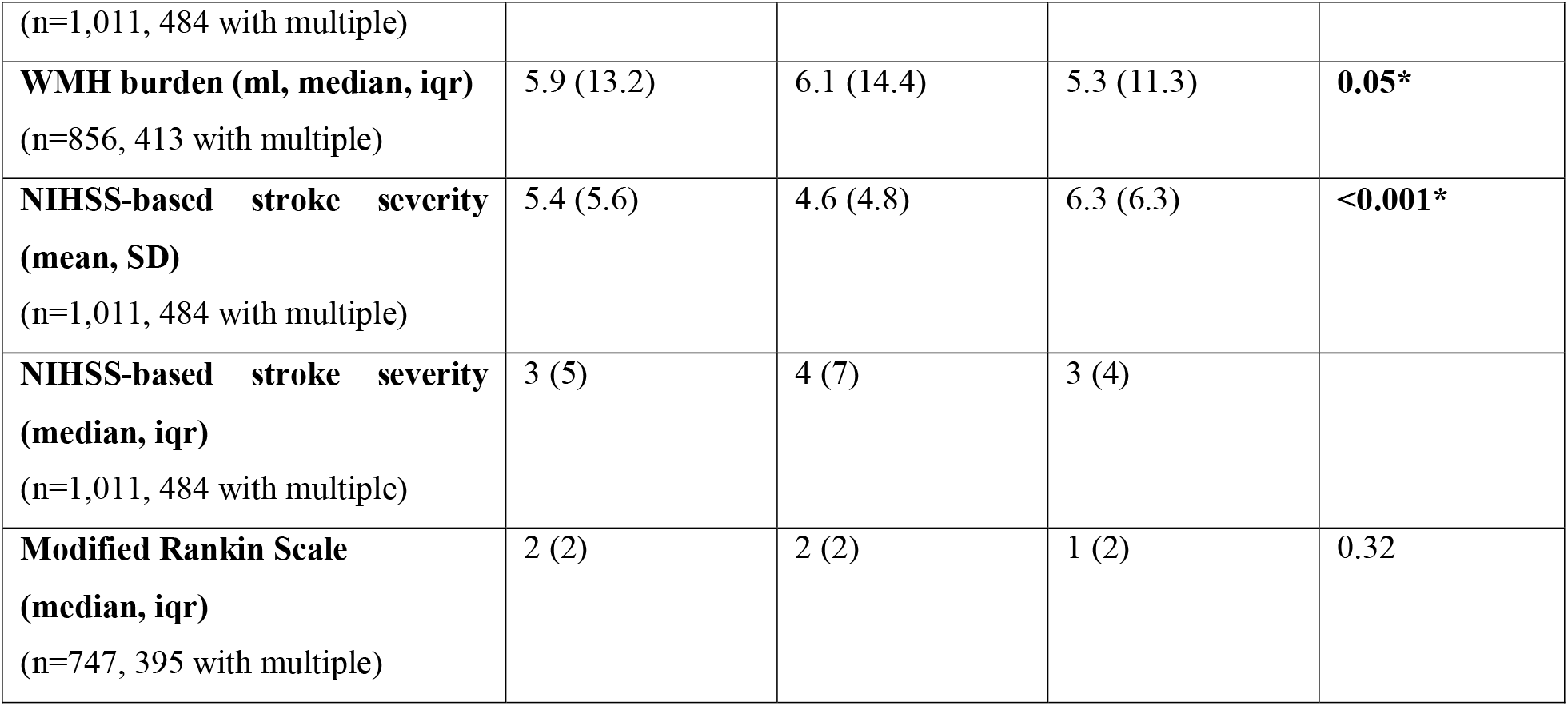
Clinical characteristics of included MRI-GENIE patients with AIS. Numbers are presented for the entire group of patients, as well as stratified based on single lesion versus multiple lesions. Statistical group differences between patients with single and multiple lesions were evaluated via two-sample t-tests or Fisher’s exact tests (level of significance p<0.05, FDR-corrected for multiple comparisons). Asterisks mark statistically significant group differences after correction for multiple comparisons.

Patients with a single ischemic lesion in a single vascular territory constituted 49.7% (1,225/2,466), while 50.3% experienced multiple ischemic lesions (1,241/2,466). Most of these patients with multiple lesions had all their lesions within one vascular territory (37.4%, 922/2,466). Further multiple lesion constellations were comparably less frequent: 4.1% (102/2,466) patients with a single lesion in a first and multiple lesions in a second vascular territory, 3.2% with two single lesions in two vascular territories. **Figure 1** presents a visual overview of these lesion and territory constellations.

**Figure 1.**
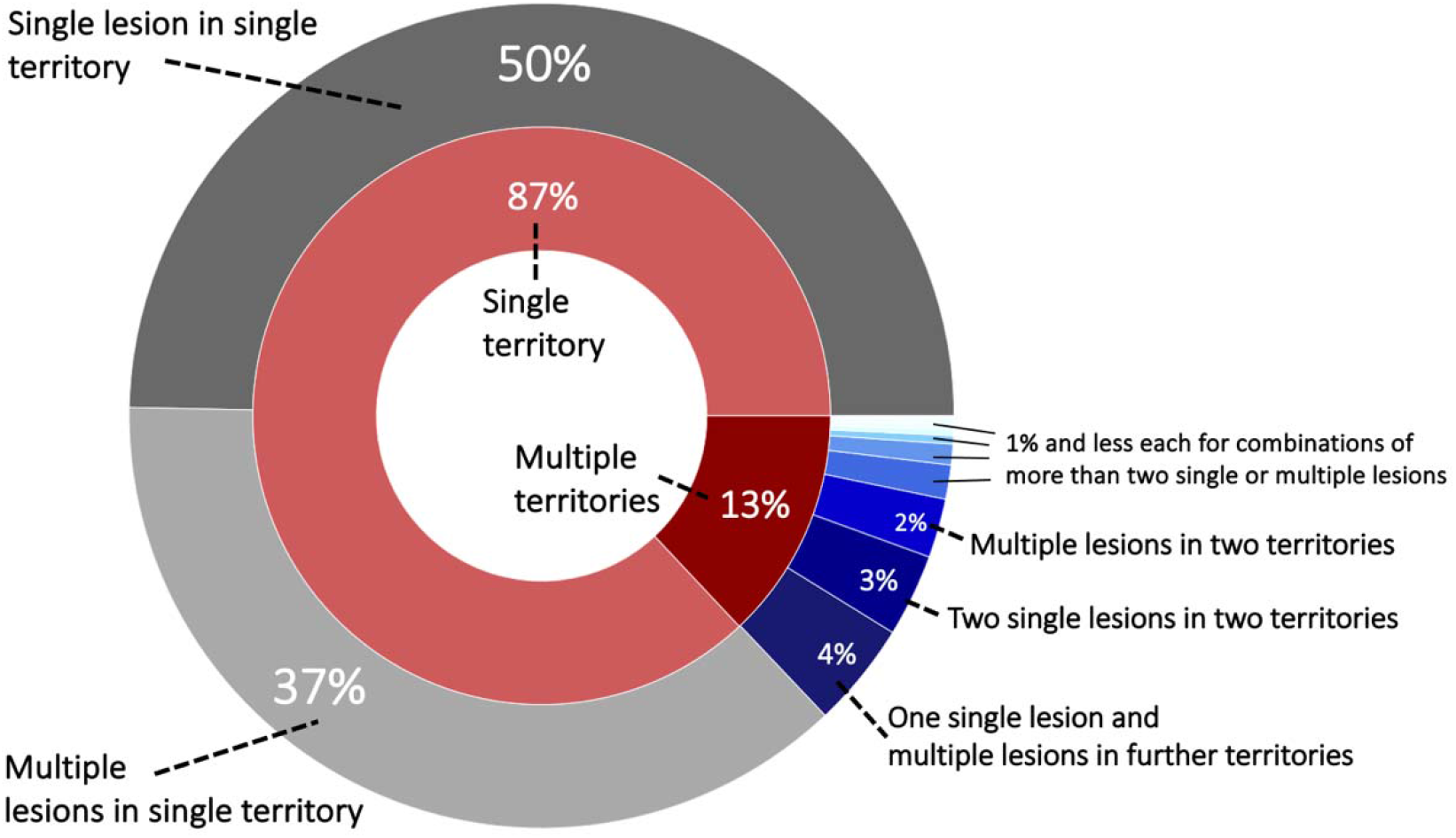
Sunburst plot of lesion and territory constellations. Most lesions, i.e., 87%, occurred within one vascular territory and the majority out of these were characterized as a single lesion (nine vascular territories: anterior cerebral artery (ACA left/right), middle cerebral artery (MCA left/right), posterior cerebral artery (PCA left/right) and vertebrobasilar territory (cerebellum left/right and brainstem)).

A total of 196 patients (8.0%) presented with bilateral stroke (ACA, MCA, PCA and cerebellar strokes, excluding the brainstem). Furthermore, 138 strokes (5.6%) occurred in both supra- and infratentorial territories and 46 (1.9%) in both the anterior and posterior circulation.

When evaluating each vascular territory separately, the frequency of multiple versus single lesions remained largely the same, i.e., ∼50% (from 43% for right ACA to 55% for right cerebellum, **Table 2**). The brainstem represented a noteworthy exception: Single lesions within the brainstem were four times more likely than multiple lesions (81% versus 19%).

**Table 2.**
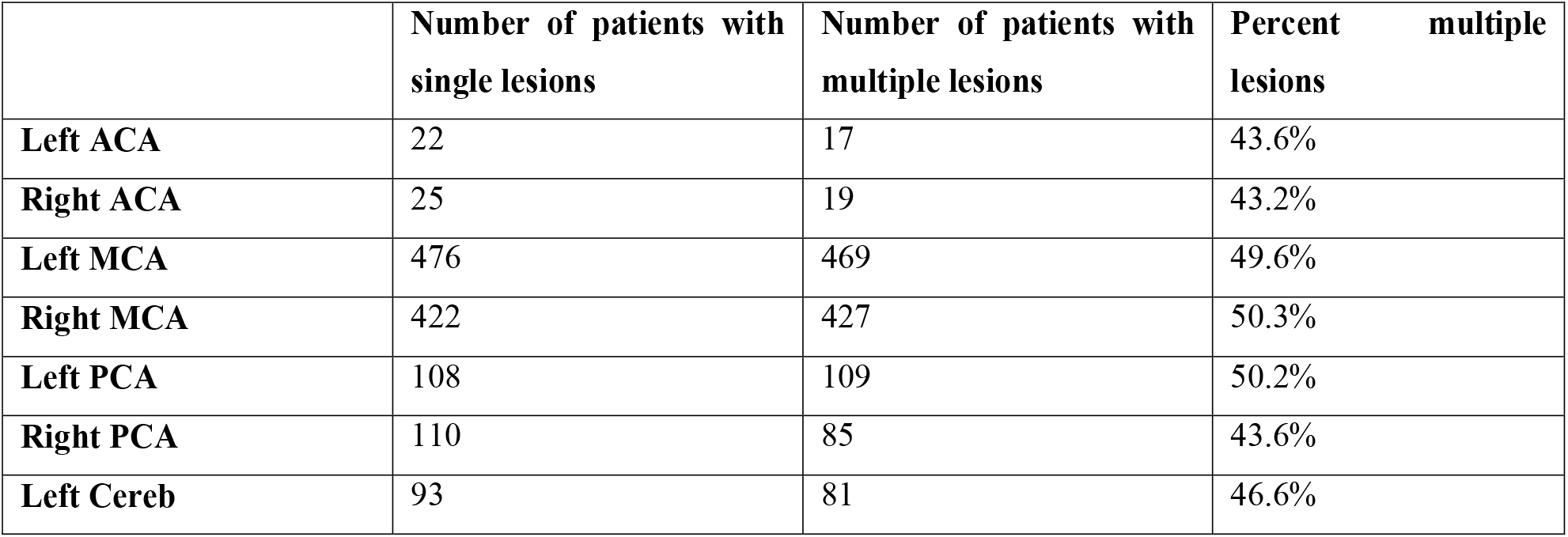
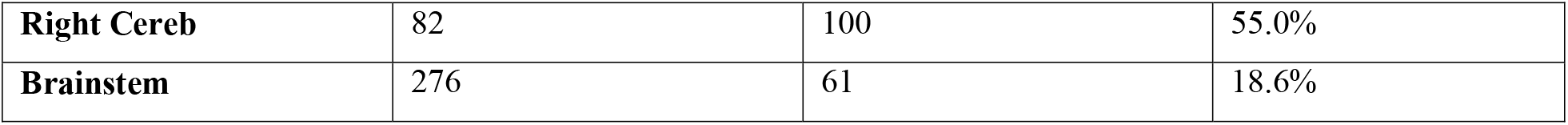
Number of patients with single and multiple lesions per vascular territory and laterality. As expected, lesions occurred most frequently in MCA territory and least frequently in ACA territory of both the left and the right hemisphere.

|  | Number of patients with single lesions | Number of patients with multiple lesions | Percent multiple lesions |
| --- | --- | --- | --- |
| **Left ACA** | 22 | 17 | 43.6% |
| **Right ACA** | 25 | 19 | 43.2% |
| **Left MCA** | 476 | 469 | 49.6% |
| **Right MCA** | 422 | 427 | 50.3% |
| **Left PCA** | 108 | 109 | 50.2% |
| **Right PCA** | 110 | 85 | 43.6% |
| **Left Cereb** | 93 | 81 | 46.6% |
| **Right Cereb** | 82 | 100 | 55.0% |
| **Brainstem** | 276 | 61 | 18.6% |

### Different clinical characteristics for patients with single and multiple lesions

There were no significant differences in age or sex between patients with a single lesion and those with multiple lesions (mean (SD) age 63.1(14.8) y, 37.7% women vs. 63.7(15.1) y, 40.2% women). Similarly, patients with single vs. multiple lesions did not differ significantly in the frequencies of the comorbidities hypertension (66.6% vs. 64.2%), diabetes mellitus (24.2% vs. 23.0%) and coronary artery disease (16.9% vs. 18.0%, all *p*-values > 0.05 after False Discovery Rate (FDR)-correction for multiple comparisons). However, significantly more patients with multiple lesions had a diagnosis of atrial fibrillation (12.4% vs. 16.9%, *p*_*FDR*_-value=0.005). Furthermore, patients with multiple lesions were more likely to be non-smokers (43.3% vs. 50.0%, *p*_*FDR*_-value=0.002). With respect to stroke etiology, stroke patients with multiple lesions were significantly more likely to be diagnosed with cardioembolic and large artery occlusion strokes (cardioembolic: 13.7% vs. 18.1%, *p*_*FDR*_-value=0.008; LAO: 16.0% vs. 30.1%, *p*_*FDR*_-value<0.001), as well as strokes of the category “other etiology” (5.7% vs. 8.6%, *p*_*FDR*_-value=0.01). In contrast, patients with single lesions more frequently experienced small artery occlusion strokes (26.0% vs. 4.0%, *p*_*FDR*_-value<0.001).

We furthermore had access to information on lesion volume and acute stroke severity for a subset of 1,011 patients (n=484 patients with multiple lesions, 47.9%). Patients with multiple lesions presented both with a higher stroke severity (NIHSS 4.6(4.8) vs. 6.3(6.3), *p*_*FDR*_-value<0.001), as well as a higher lesion volume (median (IQR) 1.7(11.2) ml vs. 7.7(30.4) ml, *p*_*FDR*_-value=0.001). Despite this higher lesion volume in the case of multiple lesions, lesion distributions themselves were qualitavely similar, featuring a predilection for subcortical infarcts in vicinity to the lateral ventricles (**Figure 2**). On the other hand, patients with single lesions were characterized by a significantly larger WMH burden (median (IQR) 6.1(14.4) ml vs. 5.3(11.3) ml, *p*_*FDR*_-value=0.048). Patients with single lesions and patients with multiple lesions did not significantly differ in their post-stroke functional outcome (median (IQR) mRS 2(2) vs. 1(2), *p*_*FDR*_-value=0.29, information available for 747 patients (n=395 patients with multiple lesions, 52.9%)).

**Figure 2.**
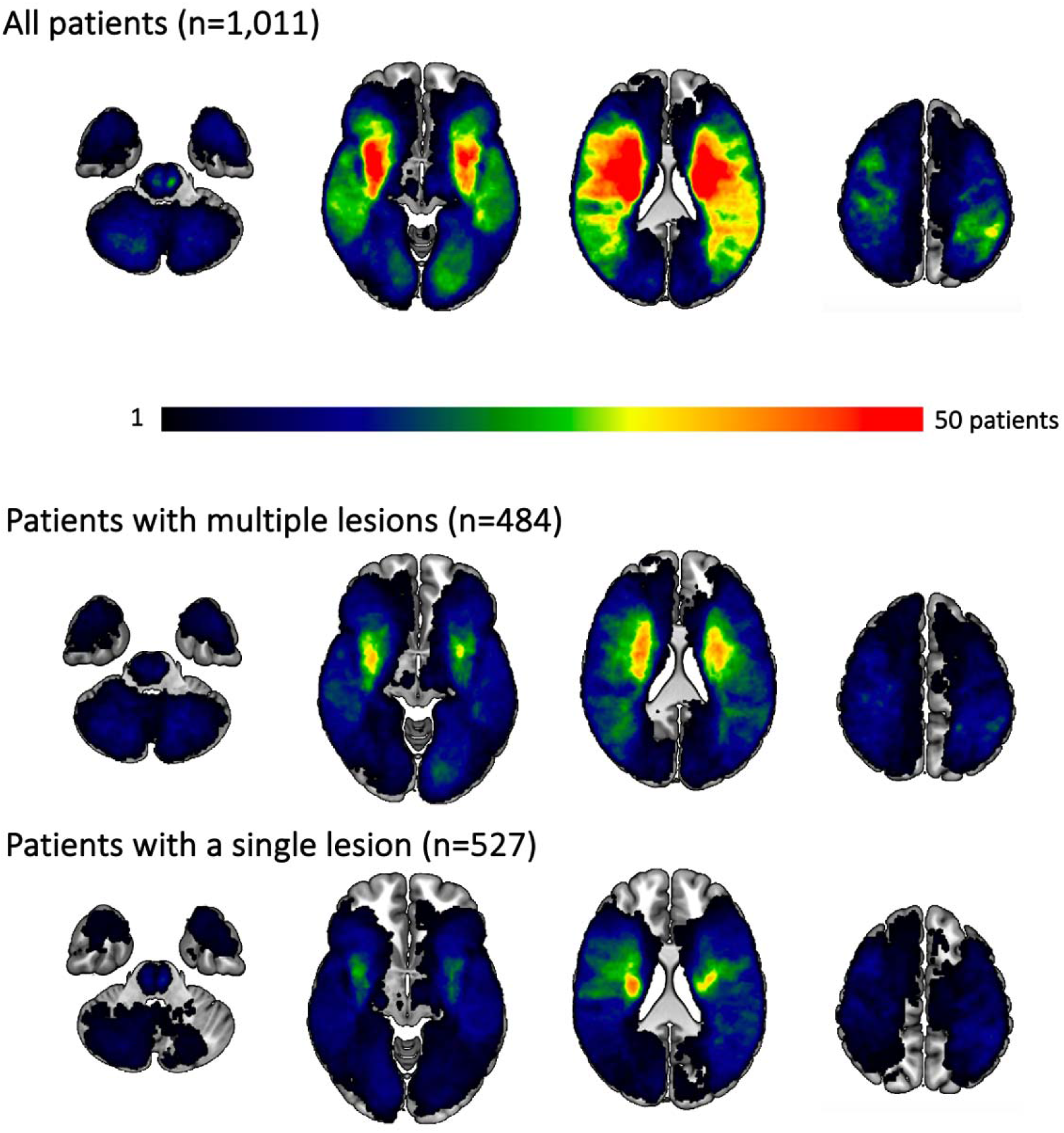
Lesion overlap for all patients and separately for those experiencing multiple and single lesions. Most lesions occurred subcortically, predominantly affecting the white matter in the vicinity of lateral ventricles. Patients with multiple lesions had larger stroke lesions on average, resulting in more extensive regions of substantial overlap. Qualitatively, the distribution of lesions was however comparable between patients with multiple and single lesions. Please note that the size of the original cohort was decreased due to lesion segmentation and stroke severity availability.

### Bayesian hierarchical regression: Interaction effects of multiple lesions and lesion volume with respect to stroke severity

These analyses were conducted within the subsample of 1,011 patients with available lesion volume and stroke severity data. Lesion volume was positively linked to stroke severity across all patients: These effects of lesion volumes were comparable between patients with single and multiple lesions (posterior distribution for single lesions: mean: 1.15, 90% HPDI: 1.01 to 1.28, posterior distribution for multiple lesions: mean: 1.21, 90% HPDI: 1.08 to 1.33; difference of posterior distributions: mean: -0.06, 90% highest probability density interval (HPDI): -0.11 to 0.003, hence overlapping with zero, **Figure 3**, upper row). Findings remained the same, when considering only those patients with anterior circulation strokes and excluding patients with lacunar lesions: Once again, the effect of lesion volume on stroke severity was comparable for patients with a single and with multiple lesions (posterior distribution for single lesions: mean: 1.56, 90% HPDI: 1.39 to 1.84, posterior distribution for multiple lesions: mean: 1.57, 90% HPDI: 1.4 to 1.76; difference of posterior distributions: mean: -0.012, 90% HDPI: - 0.091 to 0.064; **Figure 3**, middle row, c.f., **supplementary materials** for results without exclusion of lacunar stroke patients). In the case of posterior circulation stroke, lesion volume had varying effects on stroke severity depending on the single versus multiple lesion status (posterior distribution for single lesions: mean: 0.208, 90% HPDI: -0.0698 to 0.477, posterior distribution for multiple lesions: mean: 0.39, 90% HPDI: 0.127 to 0.619; difference of posterior distributions: mean: -0.182, 90% HPDI: -0.314 to -0.0689, not overlapping with zero; **Figure 3**, bottom row). Therefore, lesion volume had a more prominent role in stroke severity in the sample of patients with multiple lesions in the posterior circulation.

**Figure 3.**
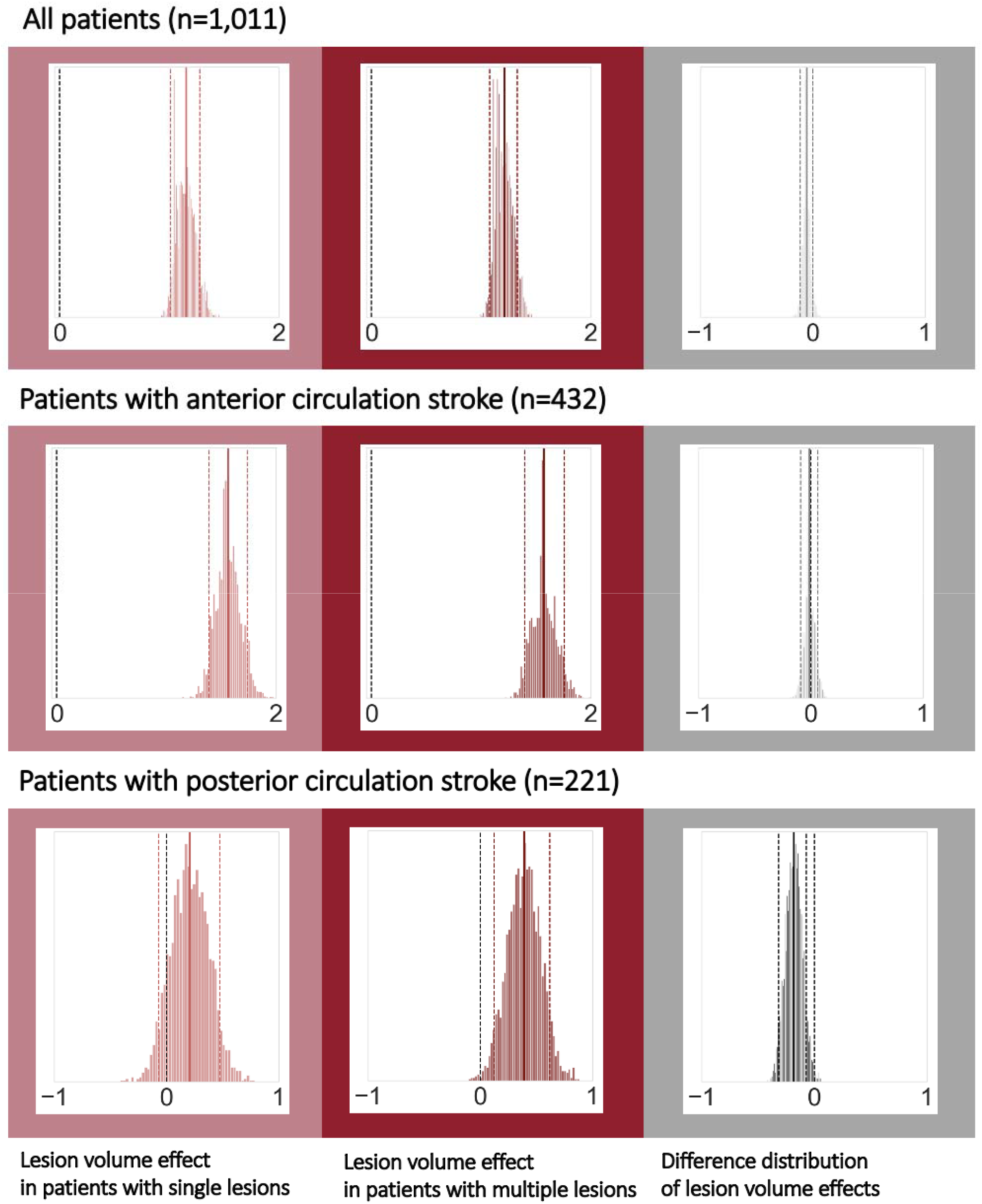
Bayesian hierarchical modeling: Lesion volume effects on stroke severity depending on the multiple versus single lesion status. The left column presents the lesion volume effects on stroke severity in the case of single lesions, while the middle column presents the lesion volume effects in the case of multiple lesions and the right column represents their difference. A total of 292 patients with non-lacunar, anterior circulation strokes had multiple lesions (hence 140 with single lesions). For posterior circulation stroke, 117 patients had multiple, and 104 patients had single lesions. Effects can be considered substantial if the zero is not included in the 90% highest probability density interval (HPDI, indicated by the dashed lines). Therefore, when analyzing all patients (upper row) or all patients with non-lacunar, anterior circulation strokes, lesion volume was noticeably linked to stroke severity – stroke severity was higher, the higher the lesion volume, independent of single or multiple lesions. However, for posterior circulation strokes, lesion volume had a substantial effect on stroke severity only in case of multiple lesions, yet not in case of single lesions. These varying links between lesion volume and stroke severity for single and multiple lesions were underscored by a difference distribution not overlapping with zero (right column, bottom row).

### Bayesian hierarchical regression: Interaction effects of multiple lesions and lesion volume with respect to functional outcomes

A total of 747 patients had available information on 3-month functional outcomes. When analyzing the entirety of patients, we once again ascertained positive links between lesion volume and unfavorable functional outcomes. These effects did not differ between the groups of patients with single and multiple lesions (difference of posterior distributions: mean: 0.002, 90% HPDI: -0.025 to 0.034). Results were qualitatively similar for patients with anterior circulation stroke and non-lacunar lesions. In case of posterior circulation stroke, lesion volume was not markedly associated with unfavorable functional outcomes, neither for single, nor for multiple lesions.

## Discussion

We leveraged a large, exceptionally well-characterized sample of 2,466 patients with AIS to investigate the intricacies of multiple ischemic lesions. We found that multiple lesions were frequent, occurring in 50% of all patients. Most of the multiple lesions occurred within one specific vascular territory; only a total of 13% of all patients had lesions in multiple vascular territories. Combined lesions in both anterior and posterior circulation brain region were rare (∼2%), contrasting with an overall higher rate of bilateral stroke lesions (∼8%). Our data corroborate previously reported higher rates of atrial fibrillation, cardioembolic and large artery occlusion etiologies^7,10,11^ and higher stroke severities^14^ in patients with MAL. Furthermore, we could gain novel insights into the links between single versus multiple lesions and the volumes of DWI, as well as WMH lesions. We found that WMH lesion volume was significantly higher in patients with single lesions. This observation can potentially be explained by the higher prevalence of small vessel etiology in single lesion stroke. It has to be noted however that the absolute difference was only 0.8ml. DWI lesion volume, in contrast, was significantly higher in those patients experiencing multiple lesions. What is more, for posterior circulation lesions, our findings suggest that lesion volume is linked to a higher stroke severity in the case of only multiple, yet not single lesions.

### Frequency of multiple lesions

Our finding that 50% of patients experience multiple lesions is in stark contrast with reports of multiple lesions in ∼2% in the earliest studies^4^ and even with those ones of up to ∼30% in more recent studies.^7^ These vastly varying numbers may arise due to both differences in the scanner technical capabilities at the time of study execution, as well as the definitions of multiple lesions. Plausibly, the sensitivity of lesion detection has substantially increased over the years due to large-scale transitions from CT to MRI scans, with MRI field strength advancing from 1.5T and 3T MRI. These changes may have resulted in even greater effects, when occurring in conjunction with thinner slice acquisition.^30^ We here only investigated MRI scans which are more sensitive than early CT scans. However, our multi-center imaging data was widely heterogeneous: Most scans were acquired at a field strength of 1.5T, some at 3T, and slice thicknesses varied from 2 to 7mm.

The increase in the percentage of patients with multiple lesions in our study may also stem from varying definitions of multiple lesions. Such differences in definitions hamper comparisons of study results in general. Our definition of “multiple” relied on whether there were several topographically discrete, isolated, unconnected lesions, independent of the actual vascular territory. This approach is in contrast to many previous studies that considered lesions to be multiple only if they occurred in several vascular territories – which, as a stricter criterion. We here, for example, observed multiple lesions in multiple territories in only ∼13% of the cases. Importantly, the exact definitions of vascular territories differed in previous studies as well: While some authors employed broad categories of hemisphere-specific anterior, middle, and posterior cerebral artery strokes (ACA, MCA and PCA, respectively),^30^ others differentiated between more subtle territories. Novotny and colleagues, for example, additionally incorporated individual leptomeningeal branches of the ACA and MCA, the anterior choroidal artery (AchA) and, in the posterior circulation, the perforating branches of the internal carotid artery, ACA, MCA, and the AchA or the vertebral arteries, basilar artery, the superior cerebellar artery, the anterior inferior cerebellar artery, the posterior inferior cerebellar artery, and the posterior cerebral artery (PCA).^12^ Baird and colleagues, on the other hand, modified criteria introduced by Bogousslavsky and collagues^31^ very early on in 1996 and discriminated between the anterior cerebral artery, middle cerebral artery (lenticulostriate, superior or inferior division), penetrating artery in the deep basal ganglia or white matter, anterior choroidal artery, and watershed strokes.^6^ Possibly, the extent of territories has an appreciable effect on the frequency of single versus multiple lesions, with fewer multiple lesions in the case of larger territories.

There may not be an objectively superior way to define multiple lesions. Practically, it may be safer to assume that lesions are truly distinct and unconnected, when occurring in separate vascular territories.^6^ However, false positives, i.e., falsely characterizing a lesion to be multiple, while it is not, may have been more of a concern, when working with low resolution (CT) images, yet less so nowadays. Altogether, it may therefore be of importance to consider the employed MAL definition when evaluating the frequency and association to clinical characteristics. In this study, we decided to employ a non-territory-based definition of multiple lesions to ascertain an upper limit for the occurrence of multiple lesions and define associations to clinical characteristics in this context, as discussed in the following.

### Multiple acute ischemic lesions and links to clinical variables

The in-depth investigations of multiple lesions are of particular clinical value if they augment our insights on potential stroke etiologies and outcomes; this new knowledge could then be instrumentalized to optimize preventative regimens and acute treatments. The high frequency of multiple lesions of 50% further underscores their relevance; many patients could potentially benefit from any optimization.

Despite differences in MAL frequency, we here observed similar constellations of stroke etiologies in patients with single vs. multiple lesions as described in previous studies:^7,10,11^ Cardioembolic and large artery occlusion etiologies were significantly more common in patients with multiple lesions, while patients with single lesions experienced significantly more small artery occlusion etiology strokes. In line with this pattern of associated etiologies, we could also ascertain a significantly higher WMH lesion load in patients with only single lesions, as well as a higher frequency of atrial fibrillation in patients with multiple lesions. However, one important observation is that these differences in stroke etiology were not absolute – any stroke etiology occurred in both single and multiple lesion stroke. For example, ∼14% of patients with a single lesion were still categorized as having had a cardioembolic stroke. In contrast, 4% of patients with multiple lesions were assigned small artery occlusion etiology. This latter finding may be well in line with prior work that described multiple lesions occurring in lacunar stroke.^9^ Importantly, the authors confirmed small artery disease as the underlying etiology in ∼57% of patients with multiple lacunar lesions. In the remaining 43% of patients with multiple lacunar lesions, etiologies other than small artery disease were determined after closer investigation. Given this detection, it may be worth re-evaluating a small artery occlusion etiology in patients with multiple lesions, especially in view of the therapeutic consequences for secondary prevention.

### Short- and long-term stroke outcomes and lesion volume

Previous studies reported varying findings with respect to the acute stroke severity, with some authors observing non-significant differences in NIHSS scores,^8,11^ while others described higher NIHSS scores, along with higher acute modified Rankin Scale scores in patients with multiple lesions.^14^ We here ascertained further evidence for a significantly higher acute stroke severity in patients with multiple lesions. Due to the availability of information on individual stroke volumes, we could enrich our investigations by additionally scrutinizing interaction effects of single versus multiple lesion status and lesion volume on outcomes. Of note, we ascertained similar links between lesion volume and stroke severity in case of anterior circulation stroke for both patients with single, as well as multiple lesions. This finding suggests that a higher stroke severity in the presence of multiple lesions may be primarily due to a correspondingly higher lesion volume. However, the nature of these links changed when focusing on patients with posterior circulation strokes. We here saw a more pronounced increase in stroke severity in case of multiple discrete lesions. In other words, the same lesion volume was linked to a higher stroke severity in case of multiple, as compared to single lesions. The brainstem, representing one of the main regions affected by a stroke in the posterior circulation, hosts numerous relevant nerve nuclei and the cortical spinal tract in close spatial proximity.

Possibly, multiple smaller lesions inevitably affect more of these centers than a larger, but single lesion could, eventually resulting in the proportionally higher stroke severity in multiple lesion stroke. Intriguingly, we observed a predominance of single lesions in the brainstem region that stood in stark contrast to all other vascular territories: While multiple lesions usually occurred in approximately half of all cases, they constituted only 19% of brainstem lesions. Future studies are warranted to explore potential explanatory mechanisms and study the effect this difference may exhibit on the effect of lesion volume on stroke outcomes.

All in all, these findings strongly support the notion that links between lesion volume and stroke outcomes need to be dissected with the greatest care possible, since the nature of associations differed depending on the single versus multiple lesion status and affected vascular territory, hence the.

Further, we did not note any significant group differences with respect to more long-term, 3-to-6-months functional outcome. These functional outcomes were measured on the modified Rankin Scale that represents a global assessment of the symptomatic consequences of stroke (0: no symptoms to 6: death).^32,33^ While the scale is most frequently employed as a primary endpoint in acute stroke treatment trials,^34,35,36^ it nonetheless captures stroke sequelae in a rather coarse-grained way. Given this limitation, it is difficult to elucidate whether the observed initial differences in stroke severity were only short-lasting and not present anymore at three months (e.g., due to a proportionally greater recovery) or were long-lasting, but too subtle to be detected. When compared to single lesions, multiple ischemic lesions were previously linked to a higher rate of death and stroke recurrence,^8^ suggesting some prolonged effect. Eventually, future investigations of long-term stroke outcomes as evaluated by more sophisticated scales, such as the Fugl-Meyer score^37^ for motor functions or the mini-mental state exam^38^ for cognitive functions, will be promising to shed further light on this matter.

### Strengths, limitations, and future evaluations

Our large sample size, as well as detailed central reads of individual scans by expert neuroradiologists are two essential strengths of this study. The multi-center character may also suggest a good generalization to stroke populations at large. However, there are multiple important limitations. It should be noted that severe stroke patients may have been underrecruited in the original studies (c.f., our median NIHSS of 3). Additionally, information on functional outcome was only available in a subset of our patient sample. Further, the focus on MRI scans may have introduced a selection bias, given that patients with CT scans were not considered. Another limitation is that we did not have readily available information on acute stroke treatment. Therefore, associations to the frequency and efficacy of thrombolysis and thrombectomy, as well as their influence on the final lesion volume remain to be estimated in future work. Similarly, it would have been interesting to follow up on previous work that highlighted links between multiple lesions and hyperviscosity,^7^ or investigated symptomatic presentations suggesting multiple lesions,^11^ but we did not have access to these clinical details. Lastly, we here focused on estimating effects for *total* DWI and WMH lesions volumes and compared stroke lesion distributions only qualitatively (**Figure 2**). Future work could go one step further even and evaluate lesion distributions and location in quantitative ways and additionally employ techniques such as radiomics to enhance the imaging-based lesion information.^39,40^

## Conclusion

Assessing a large and radiologically uniquely deep-phenotyped cohort of patients with acute ischemic stroke, we here present evidence that multiple ischemic lesions occur more frequently than previously reported, i.e., in almost half of all cases. In analyses leveraging information on lesion volume, we uncovered distinct interaction effects with multiple lesions and vascular territories: The link between lesion volume and stroke severity was the same for both single and multiple lesions in the case of anterior circulation strokes. However, in the case of posterior circulation stroke, lesion volume was linked to a higher stroke severity in multiple lesion stroke compared to single lesion stroke, but this did not carry through to ∼3-to-6-months functional outcome.

## Data Availability

Data can be made available to researchers for the purpose of reproducing the here reported results, pending the permission for data sharing by Massachusetts General Hospital's institutional review board.

## Acknowledgments

We are grateful to our colleagues at the J. Philip Kistler Stroke Research Center for valuable support and discussions. Furthermore, we are grateful to our research participants without whom this work would not have been possible.

## Funding

A.K.B. is supported by a Massachusetts General Hospital Executive Committee on Research (MGH ECOR) Fund for Medical Discovery (FMD) Clinical Research Fellowship Award. M.B. acknowledges support from the Société Française de Neuroradiologie, Société Française de Radiologie, Fondation ISITE-ULNE. A.V. is in part supported by National Institutes of Health and National Institute of Neurological Disorders and Stroke (NIH-NINDS, R01 NS103824, RF1 NS117643, R01 NS100417, U01NS100699, U01NS110772). C.J. acknowledges support from the Swedish Research Council (2018-02543 and 2021-01114), the Swedish state under the agreement between the Swedish government and the county councils, the “Avtal om Läkarutbildning och Medicinsk Forskning” (ALF) agreement (ALFGBG-720081); the Swedish Heart and Lung Foundation (20190203). A.G.L. acknowledges support from the Swedish Research Council (2019-01757), The Swedish Government (under the “Avtal om Läkarutbildning och Medicinsk Forskning, ALF”), The Swedish Heart and Lung Foundation, Region Skåne, Lund University, Skåne University Hospital, Sparbanksstiftelsen Färs och Frosta, Fremasons Lodge of Instruction Eos in Lund and National Institutes of Health (NIH, 1R01NS114045-01). N.S.R. is in part supported by National Institutes of Health and National Institute of Neurological Disorders and Stroke (NIH-NINDS, R01NS082285, R01NS086905, U19NS115388).

## Conflicts of interest

M.E. has received personal fees for consulting from Astra Zeneca and WorldCare Clinical Group. C.G. has received consulting honoraria from Microvention and Strykere and research funding from Medtronic and Penumbra. A.V. has received research funding from Cerenovus. A.G.L. has received personal fees from Bayer, Astra Zeneca, BMS Pfizer, and Portola. N.S.R. has received compensation as scientific advisory consultant from Omniox, Sanofi Genzyme and AbbVie Inc. T.U. received personal fees for consulting from AstraZeneca. All other authors declare no competing interests.

## Supplemental materials

### Methods

#### Neuroimaging parameters

Neuroimages were recorded with 1T, 1.5T or 3T scanners (General Electric Medical Systems, Philips Medical Systems, Siemens, Toshiba, Marconi Medical Systems, Picker International, Inc.).

#### Diffusion-weighted images (DWI)

Mostly axial orientation (2727/2770 axial, 43/2770 coronal). Axial: Reconstruction matrix 256×256mm^2^ (range: 128×128mm^2^ to 432×384mm^2^), median field-of-view 230 mm (range: 200 to 420 mm), median slice thickness 5mm (range: 2 to 7mm, gaps of 0 to 3mm), median TR 4.773ms, median TE 92ms. Coronal: reconstruction matrix 256×256 mm^2^, median field-of-view 260mm, median slice thickness 5mm, median TR 8.200ms, median TE 112ms. In most cases 3 directions (range: 3 to 25). In most cases low b-value 0s/mm^2^ (range: 0 to 50s/mm^2^), high b-value 1000s/mm^2^ (range: 800 to 2000s/mm^2^).

#### Model specification for stroke severity

**Hyperpriors**

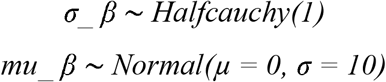

**Priors**

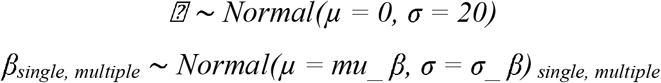

**Likelihood**

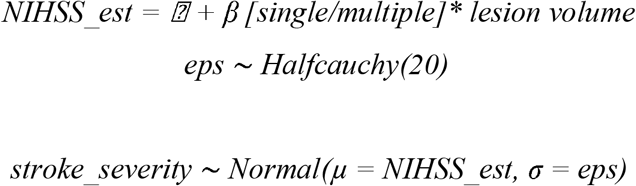

### Results

#### Bayesian hierarchical regression: Interaction effects of multiple lesions and lesion volume with respect to stroke severity

Findings remained broadly the same when analyzing patients with anterior circulation strokes and without excluding patients with lacunar lesions: Effect of lesion volume on stroke severity was similar for patients with a single and with multiple lesions (posterior distribution for single lesions: mean: 1.49, 90% HPDI: 1.33 to 1.66, posterior distribution for multiple lesions: mean: 1.51, 90% HPDI: 1.38 to 1.68; difference of posterior distributions: mean: -0.0144, 90% HDPI: -0.094 to 0.0485).

## Notes

### Author Declarations

Patients gave written informed consent in accordance with the Declaration of Helsinki. The study protocol was approved by Massachusetts General Hospital's Institutional Review Board (Protocol #: 2001P001186 and 2003P000836).

